# Multimorbidity and Medication Order Patterns in Skilled Nursing Facilities: A Real-World Evidence Study

**DOI:** 10.64898/2026.09.22.26363593

**Authors:** Huda Ashraf, Peter Tsasis, Katherine E. Mathers, Loredana Di Santo, Amy Wong, Divya Sharma, Jane M. Heffernan, Brittin Wagner, Tyler M. Saumur

## Abstract

**Background:** Multimorbidity is common among U.S. skilled nursing facilities (SNFs) residents and complicates pharmacologic management, yet real-world evidence on treatment patterns in this population remains limited. This study evaluated medication order rates and identified resident- and facility-level factors among SNF residents with multimorbidity.

**Methods:** Retrospective, observational study using electronic health record (EHR) data from PointClickCare’s Life Sciences database. Participants were 728,222 residents active in a U.S. SNF on April 30, 2025 with at least one documented diagnosis of hypertension, hyperlipidaemia, type 2 diabetes (T2D), depression, or Alzheimer’s disease and related dementias (ADRD). The outcome was presence of guideline-recommended medication order for each condition, modelled with modified Poisson regression to estimate adjusted prevalence risk ratios (RRs) and 95% confidence intervals.

**Results:** Medication order rates were 79% for ADRD, 79.1% for T2D, 87.7% for hypertension, 83.3% for hyperlipidaemia, and 90.8% for depression. Rates varied across comorbidity combinations: ADRD was associated with lower medication order prevalence, while cardiometabolic multimorbidity profiles associated with higher prevalence. Black/African American identity had the lowest likelihoods of a medication order for 3 of 5 conditions (RRs of 0.93 to 0.92), while having hypertension or hyperlipidaemia medication order was associated with higher likelihood of having orders for 3 of remaining 4 conditions (RRs of 1.02 to 1.19).

**Conclusions:** Most SNF residents with multimorbidity had a medication order for common chronic conditions, although important variability and disparities remain. Differences by comorbidity profile and social context underscore the need for targeted strategies supporting equitable, integrated prescribing and person-centred care in SNFs.

## Introduction

Multimorbidity, defined as the coexistence of two or more chronic conditions within an individual, represents a growing challenge in U.S. healthcare. ^1,2^ Approximately 50 to 80% of adults aged 65 years and older experience at least two chronic diseases and roughly 15% live with 5 or more chronic diseases. ^1,3,4^ In addition, multimorbid diagnoses are occurring at earlier ages among adults than in previous generations. ^1,5^ This trend highlights the urgent need to address the complexities associated with managing multiple chronic diseases in diverse patient populations.

The burden of multimorbidity is substantial both clinically and economically. Multimorbidity reflects a dynamic and interconnected system in which biological, psychological, social, and economic factors interact. From a complexity theory perspective, health systems and patient experiences can be understood as complex adaptive systems, whereby outcomes emerge from the interactions among patients, healthcare providers, institutions, policies, and broader social determinants of health. ^6^ Clinically, coexisting conditions contribute to frailty, reduced physical functioning, increased risk for serious/complex mental health challenges, and higher mortality risk among older adults. ^7-9^ Economically, healthcare costs and out-of-pocket expenses are both negatively impacted by multimorbidity, ^10,11^ supporting the likelihood of cost-related nonadherence, whereby individuals skip appropriate use of medication or delay receiving healthcare due to economic barriers. ^12^ Within the U.S., the burden of multimorbidity is further accentuated by social disadvantage, with individuals living in under-resourced communities exhibiting 17.1% to 18% higher initial chronic disease counts and higher rates of end-stage organ damage related to their multimorbidity state than those in the least deprived areas. ^13,14^ Furthermore, people with multiple chronic conditions are often underserved by care systems designed for single diseases, highlighting significant challenges for equity and health-system performance. ^15^

Increasing comorbidity is associated with greater treatment burden, including more complex medication regimens and a higher risk of potential drug-drug and drug-disease interactions. ^16^ For example, although statins effectively reduce low-density lipoprotein cholesterol in many populations, this benefit is not consistently observed among individuals with end-stage kidney disease. ^17^ Similarly, non-selective beta blockers are contraindicated in patients with asthma, ^18^ despite asthma and heart failure frequently co-occurring as comorbid conditions. ^19^ Conversely, some medications may have therapeutic benefits across multiple conditions; for instance, selective serotonin reuptake inhibitors are commonly prescribed to treat depression and anxiety and may also play a role in managing symptoms associated with Alzheimer’s disease. ^20,21^ Together, these examples illustrate the complex interplay among coexisting diseases, symptom profiles, and pharmacologic mechanisms, which further complicate treatment decision-making for individuals with multimorbidity.

This interplay raises critical questions: 1) Which combinations of chronic conditions are least likely to receive appropriate pharmacologic therapy? 2) Which social, demographic, or facility factors primarily contribute to undertreatment? And 3) How can prescribing practices in skilled nursing facility (SNF) settings be optimized to ensure equitable access to treatment? Using a large U.S. SNF dataset, this study aims to evaluate medication order patterns among older adults with multimorbidity for 5 patient cohorts consisting of: Alzheimer’s disease and related dementia (ADRD), type 2 diabetes (T2D), hypertension, hyperlipidaemia, and depression.

## Methods

### Setting and Participants

This retrospective observational study utilized deidentified electronic health record (EHR) data from the PointClickCare Life Sciences database, one of the largest real-world datasets of its kind, encompassing clinical information from more than 18 million residents across U.S. assisted living facilities, senior living, and skilled nursing facilities (SNFs) from 2015 to present. The database offers a comprehensive view of prescribing practices, comorbidity patterns, and resident demographics in the long-term care population. Resident data were deidentified with expert-determined approval in accordance with privacy standards and consent was obtained through business associate agreements with long-term care facilities. Exemption from ethics committee oversight was granted by the Research Ethics Boards of the University of Toronto and University of Alberta.

Data were assessed cross-sectionally using a database snapshot of April 30, 2025, with eligible residents required to be active in a skilled nursing facility on that date. EHR data were available for 787,292 individuals, of which 728,222 met inclusion criteria for this study. Residents were required to have at least one active documented diagnosis of hypertension, hyperlipidaemia, T2D, depression, or ADRD on the database snapshot date.

### Exposures and Variables of Interest

Residents were categorized based on the presence or absence of pharmacologic therapy orders consistent with clinical practice guidelines. Variables included demographic characteristics (age, sex, race/ethnicity), insurance status, facility region, and length of stay. Clinical characteristics were assessed using the Charlson Comorbidity Index score (as coring system that predicts a patient’s 1-year mortality risk and long-term outcomes), active ICD-10 codes, and counts of unique over-the-counter and prescription medications. To understand organizational and environmental influences, facility-level variables were included. These consisted of Social Vulnerability Index (SVI) scores, facility size, and facility type. SVI scores were based on demographic and socioeconomic data collected by the Centers for Disease Control and Prevention on the facility’s geographic region. ^22^

### Data Analysis

Data were extracted using SQL and analysed in Python. Descriptive statistics were used to summarize resident demographics, comorbidity distributions, and medication use.

Medication order rates were calculated for each treatment class (ADRD medications, T2D medications, depression medications, hypertension medications, and hyperlipidaemia medications) for each multimorbidity profile. A modified Poisson regression model was used with a log link to estimate prevalence risk ratios (RRs) and 95% confidence intervals (CIs) for having a medication order. Covariates included resident demographics, clinical characteristics, medication/diagnosis count, and facility characteristics while categorical variables were dummy-coded with pre-specified reference categories: female sex, White race, Medicare fee-for-service payer, Southern U.S. region, and non-profit facility ownership. Continuous variables were rescaled prior to model entry to improve interpretability of effect estimates: age was expressed per 10-year increment, BMI per 5- unit increment, number of facility beds per 25-bed increment, and days in facility per 30- day increment . Interactions were limited to 2-way comorbidity interactions to minimize multicollinearity. Statistical significance was defined as p <0.001.

## Results

### Demographic and Clinical Characteristics

The study population was predominantly within the 70s age range (mean [SD] age of 76.5 [12.6] years), female (59.2%), and White (66.9%; see Table 1). Residents were represented across all U.S. Census regions, with the largest proportions located in the South (36.4%) and Midwest (28.3%). Medicaid was the most common primary payer (32.9%), followed by Medicare Fee-for-Service (28.2%) and Managed Care (24.0%).

**Table 1.** Demographic and Clinical Characteristics.

|  | **N Residents**<br>**728,222** |
| --- | --- |
| **Age Category, n (%)** |  |
| ≥90 | 111,387 (15.3%) |
| 85-89 | 98,554 (13.5%) |
| 80-84 | 110,365 (15.2%) |
| 75-79 | 111,546 (15.3%) |
| 70-74 | 97,706 (13.4%) |
| 65-69 | 79,773 (11.0%) |
| 19-64 | 118,891 (16.3%) |
| **Age, years** |  |
| Mean (SD) | 76.5 (12.6) |
| Median (Q1, Q3) | 78 (69, 86) |
| Min, Max | 20, 90 +<sup>a</sup> |
| **Sex, n (%)** |  |
| Female | 431,020 (59.2%) |
| Male | 297,202 (40.8%) |
| **Race, n (%)** |  |
| White | 487,419 (66.9%) |
| Black or African American | 116,795 (16.0%) |
| Unspecified | 85,091 (11.7%) |
| Asian | 15,506 (2.1%) |
| Hispanic or Latino | 14,918 (2.0%) |
| American Indian or Alaska Native | 7,068 (1.0%) |
| Native Hawaiian or Other Pacific Islander | 1,425 (0.2%) |
| **Region, n (%)** |  |
| South | 264,969 (36.4%) |
| Midwest | 206,142 (28.3%) |
| Northeast | 146,009 (20.1%) |
| West | 111,102 (15.3%) |
| **Primary Payer, n (%)** |  |
| Medicaid | 239,258 (32.9%) |
| Medicare FFS | 205,056 (28.2%) |
| Managed Care | 174,878 (24.0%) |
| Private | 82,843 (11.4%) |
| Other | 26,187 (3.6%) |
| **CMS Overall Rating** |  |
| Mean (SD) | 2.6 (1.4) |
| Median (Q1, Q3) | 2 (1, 4) |
| Min, Max | 1, 5 |

**Table 1. Demographic and Clinical Characteristics**
|  |  |
| --- | --- |
| **SVI Overall Percentile** |  |
| Mean (SD) | 0.7 (0.2) |
| Median (Q1, Q3) | 0.8 (0.6, 0.9) |
| Min, Max | 0.0, 1.0 |
| **Number of Beds** |  |
| Mean (SD) | 134.2 (73.0) |
| Median (Q1, Q3) | 120 (95, 155) |
| Min, Max | 4, 815 |
| **Days in Facility** |  |
| Mean (SD) | 686.7 (786.4) |
| Median (Q1, Q3) | 398 (88, 1,006) |
| Min, Max | 2, 3,804 |
| **Stay Type, n (%)** |  |
| Long Stay | 535,796 (73.6%) |
| Short Stay | 192,426 (26.4%) |
| **Condition, n (%)** |  |
| Hypertension | 553,519 (70.3%) |
| Hyperlipidemia | 447,080 (56.8%) |
| Type 2 Diabetes | 303,680 (38.6%) |
| ADRD | 295,946 (37.6%) |
| Depression | 255,892 (32.5%) |
| **CCI Score** |  |
| Mean (SD) | 2.8 (2.2) |
| Median (Q1, Q3) | 2 (1, 4) |
| Min, Max | 0, 23 |
| **Prescription Medication Count** |  |
| Mean (SD) | 9.2 (4.7) |
| Median (Q1, Q3) | 9 (6, 12) |
| Min, Max | 0, 73 |
| **Over-The-Counter Medication Count** |  |
| Mean (SD) | 6.9 (3.7) |
| Median (Q1, Q3) | 7 (4, 9) |
| Min, Max | 0, 44 |
| **Active Diagnosis Count<sup>b</sup>** |  |
| Mean (SD) | 22.0 (10.8) |
| Median (Q1, Q3) | 20 (15, 27) |
| Min, Max | 0, 247 |

Abbreviations: ADRD=Alzheimer's disease and related dementias; CCI=Charlson Comorbidity Index; FFS=fee-for-service; ICD=International Classification of Diseases; Q=quartile; SD=standard deviation.
<sup>a</sup> The maximum age range was capped at 90+ to protect participant privacy, as individuals above 90 years represent a very small subgroup and may be more readily identifiable.
<sup>b</sup> Active diagnoses were based on ICD-10 codes.

The overall cohort was marked by substantial clinical complexity and extended residence in SNF settings. Approximately three-quarters of residents (73.6%) were classified as long-stay, with an average length of stay of ∼2 years. Hypertension was the most prevalent condition of interest, affecting 70.3% of residents, followed by hyperlipidaemia (56.8%), T2D (38.6%), ADRD (37.6%), and depression (32.5%). Residents had a mean (SD) of 22.0 (10.8) active diagnoses and had a mean (SD) Charlson Comorbidity Index score of 2.8 (2.2), which is a composite measure reflecting the cumulative burden and severity of coexisting conditions. This clinical complexity was mirrored in residents’ medication order--the mean (SD) number of prescription and over-the-counter medications was 9.2 (4.7) and 6.9 (3.7), respectively.

### Multimorbidity Profiles

Within the ADRD cohort, the most common comorbid combinations were ADRD+HLD+HTN (15.6%), ADRD+HLD+HTN+T2D (11.8%), and ADRD+HTN (11.8%; Figure 1). ADRD+T2D+MDD was the least prevalent ADRD combination, occurring in just 0.8% of residents. The overall medication order rate for ADRD was 79%. Medication order patterns differed substantially across the multimorbidity profiles. Higher ADRD medication order rates were observed among residents with depression and lower rates were typically more common with T2D as a comorbidity.

**Figure 1.**
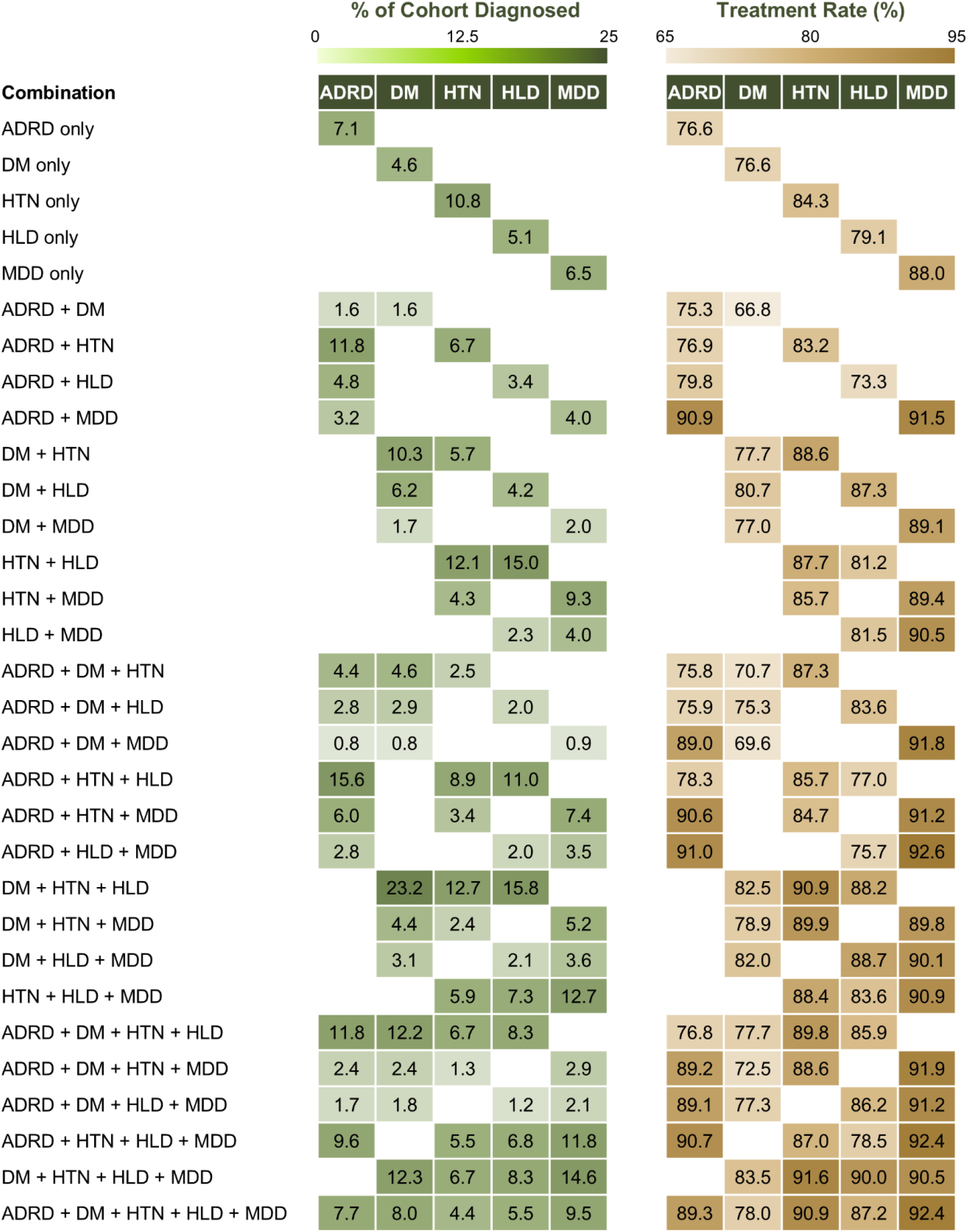
Medication order rates across multimorbidity profiles. The heatmaps display the proportion of residents with a medication order for each condition-specific medication class across multimorbidity profiles. Rows represent combinations of comorbid conditions and columns represent medication classes. Cell values indicate medication order rates (%) among residents within each multimorbidity profile.

Within the T2D cohort, the most common comorbid combinations were T2D+HTN+HLD (23.2%), T2D+HTN+HLD+MDD (12.3%), and ADRD+T2D+HTN+HLD (12.2%).

ADRD+T2D+MDD was the least prevalent combination, occurring in 0.8% of residents. The overall medication order rat e for T2D was 79.1%. Higher T2D treatment rates were observed among residents with hyperlipidaemia . In contrast, substantially lower treatment rates were observed among residents with ADRD.

Within the hypertension cohort, the most prevalent comorbid combinations were T2D+HTN+HLD (12.7%), HTN+HLD (12.1%), and HTN only (10.8%). ADRD+T2D+HTN+MDD was the least prevalent combination, occurring in 1.3% of residents. The overall prevalence of medication orders for hypertension was higher than the other conditions at 87.7%. Higher medication order rates were observed among residents with concurrent T2D and hyperlipidaemia. Conversely, lower hypertension medication order rates were observed among residents with ADRD+HTN (83.2%), HTN only (84.3%), and ADRD+HTN+MDD (84.7%).

The most frequently observed comorbid combinations in the hyperlipidaemia cohort were T2D+HTN+HLD (15.8%), HTN+HLD (15%), and ADRD+HTN+HLD (11%).

ADRD+T2D+HLD+MDD was the least prevalent combination, occurring in 1.2% of residents. The overall medication order rate for hyperlipidaemia was 83.3%. A higher prevalence of medication orders was observed with the T2D comorbidity present. Conversely, a lower prevalence was observed among ADRD comorbidity combinations.

The most common comorbid combinations in the depression cohort were T2D+HTN+HLD+MDD (14.6%), HTN+HLD+MDD (12.7%), and ADRD+HTN+HLD+MDD (11.8%).

ADRD+T2D+MDD was the least prevalent combination, occurring in 0.9% of residents. The overall prevalence of medication orders for depression was the highest among conditions at 90.8%. Higher rates of medication orders were consistently associated with ADRD as a comorbidity. Lower rates were observed among residents with MDD only (88%), T2D+MDD (89.1%), and HTN+MDD ( 89.4%).

#### Risk Ratios

Risk ratios (RRs) were estimated using modified Poisson regression models to examine predictors of pharmacologic treatment receipt across all five conditions, with RRs ranging from 0.87 to 1.19 (Supplementary Tables 1 –5). Figure 2 highlights the five highest and lowest statistically significant risk ratios for each condition. Several key patterns can be observed across conditions. Similar to the comorbidity findings, hypertension (RR range: 1.02-1.16) and hyperlipidaemia medication orders (RR range: 1.04-1.19) were among the strongest positive predictors of medication orders for the remaining conditions, excluding ADRD. Black or African American race was among the strongest positive predictors of medication orders for hypertension and hyperlipidaemia, with modest RRs [95% CIs] of 1.05 [1.04, 1.05] and 1.04 [1.04, 1.04], respectively. Conversely, it was among the strongest negative predictors of medication orders for ADRD (RR=0.93 [0.93, 0.94]), depression (RR=0.95 [0.95, 0.95]), and T2D (RR=0.97 [0.96, 0.97]). Residents of Asian race had a higher likelihood of receiving medication orders for T2D (RR=1.06 [1.05, 1.07]) and hyperlipidaemia (RR=1.06 [1.05, 1.07]), but a lower likelihood of ADRD medication orders (RR = 0.93 [0.92, 0.95]). Having a primary payer type of “other” was associated with a lower likelihood for medication orders, except ADRD with an RR range of 0.87-0.97. Lastly, an active ADRD medication order was the strongest predictor of having a depression medication order (RR=1.04 [1.04, 1.05]) and vice-versa, although depression medication was a much stronger predictor (RR=1.14 [1.13, 1.15]).

**Figure 2.**
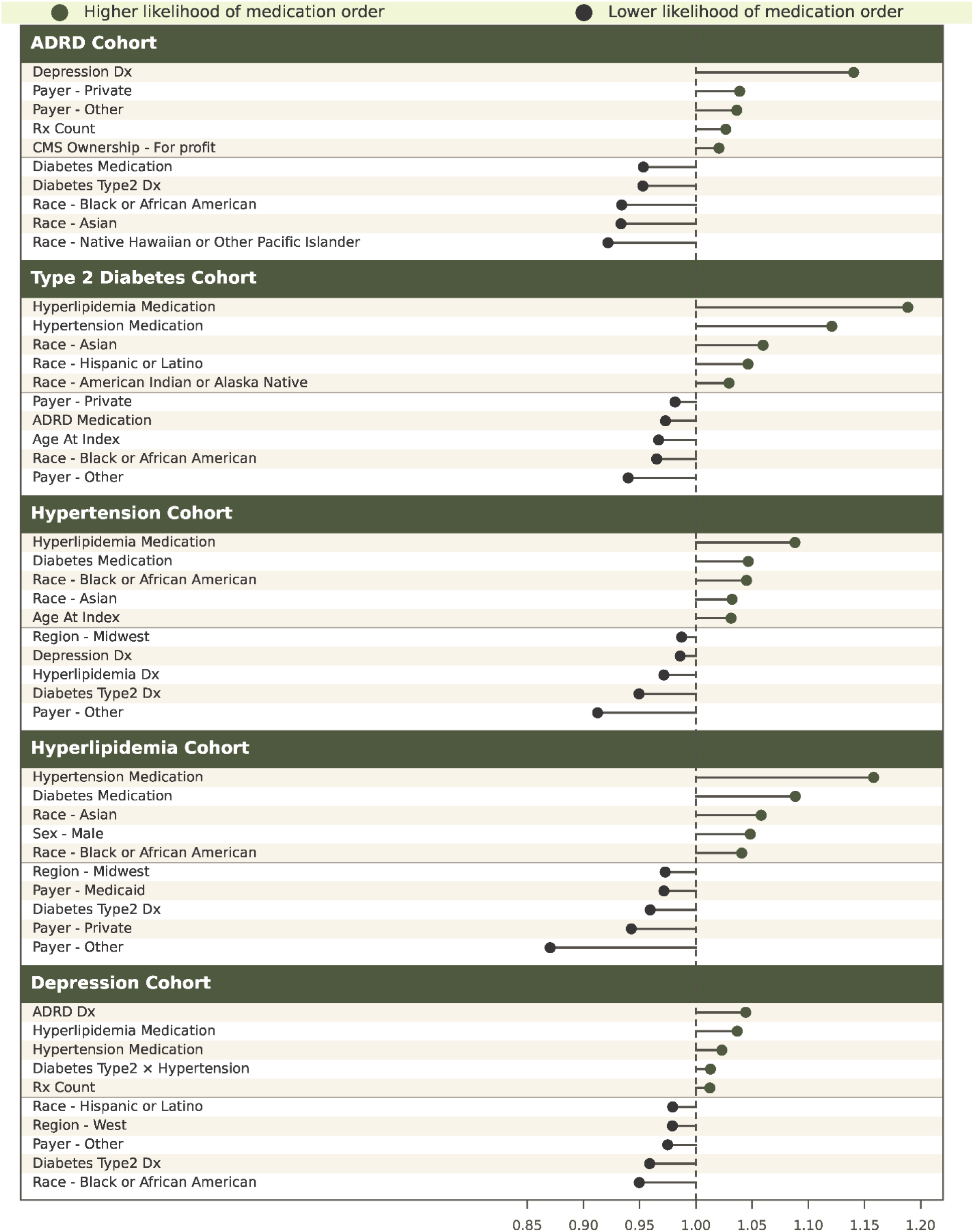
Summary of 5 highest and 5 lowest risk ratios. Abbreviations: ADRD=Alzheimer’s disease and related dementias; CMS=Centers for Medicare and Medicaid; Dx=diagnosis; Rx=prescription; Type2=type 2 diabetes; ×=interaction term. Note: The figure presents variables in the model with risk ratios with a p<0.001 .

## Discussion

In this large national study of United States SNF residents, we observed substantial heterogeneity in medication order patterns among individuals with multimorbidity, underscoring the complexity of managing multiple chronic conditions in institutional settings. Although the prevalence of medication orders for ADRD, T2D, hypertension, hyperlipidaemia, and depression were generally high (>75% overall), important gaps remained. Treatment receipt varied meaningfully across multimorbidity profiles, suggesting that prescribing decisions in SNF may reflect prioritization of certain conditions over others rather than integrated management of coexisting diseases. ^10,15^

Firstly, it was found that cardiometabolic conditions remain strongly prioritized in SNF as medication orders for cardiometabolic conditions appeared more consistent across multimorbidity profiles. Residents with all three cardiometabolic disorders (T2D+hyperlipidemia+hypertension) formed a large cohort of 70,502 residents and demonstrated the highest prevalence of medication orders for T2D, hyperlipidaemia, and hypertension medications. Similarly, residents with depression in combination with these cardiometabolic conditions also showed the highest medication order rates of antidepressant medication. These patterns suggest that cardiometabolic conditions may remain strongly prioritized in SNFs due to established prescribing pathways and clearer short-term clinical targets as prior analyses of Medicare beneficiaries similarly show that individuals with high multimorbidity often continue to receive aggressive cardiometabolic treatment despite increasing functional impairment and care complexity. ^10^

Further, while residents with T2D, hyperlipidaemia, and hypertension generally had a higher prevalence of medication orders, the presence of ADRD as a comorbid condition was associated with lower medication order rates across most profiles. This suggests that prescribing patterns may differ when dementia coexists with multiple cardiometabolic conditions. Furthermore, those in SNF facilities are often in later stages of dementia and have either previously taken medications such as cholinesterase inhibitors or are at a stage where there would be limited benefit. Thus, the overall lower prevalence of medication orders for ADRD may reflect appropriate deprescribing patterns, rather than undertreatment. The strong association between ADRD and depression treatment in the present study likely reflects the substantial overlap of neuropsychiatric symptoms and shared clinical management approaches in SNF populations. In SNF settings, where medication burden and clinical complexity are substantial, clinicians may be particularly cautious about initiating or continuing dementia-related therapies alongside multiple cardiometabolic agents.

Medication order patterns also varied by payer type and race/ethnicity. Several minority racial groups demonstrated lower likelihoods of medication orders for specific conditions, particularly ADRD and depression, suggesting that disparities in chronic disease management remain evident even within institutional SNF settings. Prior research has shown that socioeconomically disadvantaged populations typically exhibit poorer outcomes, ^22,23^ which may result in greater prioritization for treatment once admitted into long-term care. These findings reinforcing existing evidence that social disadvantage intensifies both the burden and management challenges of multimorbidity in the United States. ^24^ The racial disparities observed in this study related to depression medication particularly in the Black/African American community are well documented and often associated with stigma and a lack of confidence and trust in mental health care. ^25,26^ While approaches to combat these barriers such as psychoeducation and eliminating microaggressions during therapy have been suggested, ^27^ our findings appear to suggest that these disparities in treatment are still present.

### Implications for policy, practice and research

Taken together, these findings highlight the tension between disease-specific prescribing and holistic care for residents with multimorbidity. The observed differences in treatment for dementia and cardiometabolic conditions suggest that clinical prioritization, medication burden, and social context jointly shape prescribing decisions in SNFs. Future research should focus on distinguishing appropriate clinical non treatment from remediable gaps in care and on developing pragmatic strategies, such as structured medication reviews, that can reduce unwarranted variability without increasing polypharmacy or treatment burden.^28^

From a practice and policy perspective, these results highlight the need for strategies that support integrated, person-centred prescribing for residents with multimorbidity while minimizing unnecessary polypharmacy. Structured medication reviews, enhanced clinical decision support, and greater training around cultural differences and views regarding pharmacological treatment may help optimize treatment options for individuals with comorbidities.

### Strengths and limitations

This study has several strengths, including its scale and national scope, drawing on more than 700,000 residents from one of the largest real-world long-term care EHR datasets in the United States, and its characterization of prescribing across concurrent condition combinations rather than one disease at a time. Several limitations should be considered when interpreting these findings. Treatment was defined based on medication orders documented in the electronic health record and does not capture medication administration, adherence, duration, or clinical appropriateness. The relatively short observation window limits assessment of longitudinal prescribing trajectories, including deprescribing or treatment escalation over time. Diagnoses were identified using routine diagnostic coding and may be nonspecific or incomplete. Finally, although this study draws from a large national dataset of United States SNFs, findings may not generalize to other care settings or health systems. Ongoing challenges in accurately measuring medication exposure in nursing homes further highlight the need for cautious interpretation. ^29^

## Conclusions

The findings of this national EHR study demonstrate substantial variability in the delivery of guideline directed pharmacologic therapy among SNF residents with multimorbidity, despite generally high overall treatment rates across conditions. Treatment receipt differed meaningfully by comorbidity profiles, with cardiometabolic conditions consistently prioritized and more variable treatment observed for Alzheimer disease and related dementias.

Overall, these findings suggest that pharmacologic management in multimorbidity is not guided by a unified, patient-centred approach to coexisting disease burden, but rather by selective prioritization of individual conditions based on perceived clinical urgency and treatment familiarity. Treatment variability across multimorbidity profiles further highlights how social determinants of health and contextual factors shape access to and delivery of care, reinforcing the need for more integrated and equitable approaches to medication management in long-term care populations.

## Data Availability

The data underlying this study are proprietary to PointClickCare Life Sciences and were used under a business associate agreement with participating long-term care facilities. Restrictions apply to their availability; they are not publicly available. Requests for access may be directed to the corresponding author and are subject to PointClickCare Life Sciences' approval.

## Acknowledgements

We would like to thank Michelle Sweeny and Aaron Norfolk for providing their perspectives and input. This research was supported by PointClickCare Life Sciences, McMaster University, and York University. McMaster University Library provided journal article access.

